# Longitudinal Brain Correlates of Cognitive Performance in Early Psychosis

**DOI:** 10.64898/2026.08.28.26361680

**Authors:** Kossivi Armel Mignondje, Jillian G. Connolly, Adam Beermann, Ella Crabtree, Simon Vandekar, Maxwell Roeske, Kathryn Biernacki, Michael J. Coleman, Martha E. Shenton, Roscoe O. Brady, Kathryn E. Lewandowski, Heather Burrell Ward

**Author notes:** Authors Contributed Equally (Kossivi Armel Mignondje, Jillian G. Connolly, Kathryn E. Lewandowski, Heather Burrell Ward). <u>Corresponding Author:</u> Heather Burrell Ward, MD, Department of Psychiatry and Behavioral Sciences, Vanderbilt University Medical Center, 1601 23^rd^ Ave S, Nashville, TN 37212.

## Abstract

**Background:** Cognitive impairment is the leading cause of disability in schizophrenia with limited treatments. A major barrier to treatment development is the absence of reproducible, mechanistically grounded neural targets. Cross-sectional studies have identified dorsomedial prefrontal cortex (DMPFC)-somatomotor connectivity as a neural marker of cognitive performance on the Auditory Continuous performance task (ACPT), a measure of attention. To test the stability of this marker, we tested the relationship between DMPFC-somatomotor connectivity and ACPT performance in a longitudinal psychosis sample.

**Methods:** Individuals with early psychosis (n=251) and matched controls (n=90) were enrolled and underwent resting-state neuroimaging and neurocognitive assessment. A subset completed longitudinal assessments over 2-4 years. We calculated DMPFC-somatomotor resting-state functional connectivity using a previously identified DMPFC region and a seed in the somatomotor cortex. We performed linear mixed effects models to predict ACPT performance based on connectivity, time, psychosis type, and their interaction.

**Results:** In the psychosis sample, time (p=.0037) and affective psychosis diagnosis (p<.0001) predicted better ACPT performance. In a model predicting ACPT performance, we observed a significant interaction effect of DMPFC-somatomotor connectivity*psychosis subtype (p=.0079) such that DMPFC-somatomotor connectivity predicted ACPT performance only in individuals with non-affective psychosis (p=.0051). We then tested the specificity of this connectivity-cognitive performance relationship. In a model predicting DMPFC-somatomotor connectivity, only ACPT performance (p=.017), but not fluid cognition, was a significant predictor.

**Conclusions:** DMPFC-somatomotor connectivity is longitudinally associated with cognitive performance in early psychosis. This relationship is strongest in nonaffective psychosis, suggesting a novel, reliable target for intervention for cognitive deficits in early psychosis.

## Introduction

Cognitive deficits are a leading cause of functional impairment in schizophrenia and contribute to difficulties in daily functioning (1). Individuals with schizophrenia are less likely to maintain long-term employment, enter higher education, and sustain long-term social relationships compared to non-psychosis populations (2–4). Specifically, deficits in working memory, attention, and executive function in schizophrenia have been reliably identified as central impairments which may contribute to poor functional outcomes (5). Unlike positive and negative symptoms, cognitive deficits are often present before the onset of psychotic symptoms and steadily persist for the duration of illness (6). This distinction makes early psychosis a unique timepoint where interventions to prevent cognitive decline may be most effective (7–11).

Despite the known detrimental effects caused by neurocognitive deficits in psychotic disorders, there are currently no pharmacologic treatments for cognitive impairment (12). Treatment options for cognitive impairment in psychosis may be inadequate due to the limited understanding of the neural circuitry underlying the cognitive dysfunction (13). Although single studies have identified neural circuits linked to cognitive performance, reliable and reproducible neural correlates of cognitive function have not been identified (14,15).

In order to develop novel, circuit-based therapeutics for cognitive deficits in psychotic disorders, we need to identify neural markers of cognitive performance that can be modulated. A promising neural marker would need to be readily and reproducibly identifiable, exist across a population of people with psychosis, and be reliably linked to cognitive deficits within individuals across time.

In previous work attempting to address this gap, we identified a reproducible association between dorsomedial prefrontal cortex (DMPFC)-somatomotor connectivity and cognitive performance in psychotic disorders using the Seidman Auditory Continuous Performance Task (ACPT) across two independent psychosis spectrum populations (16). The ACPT is a performance-based task specifically designed to assess sustained attention in individuals with psychosis or at risk for psychosis, and it is particularly sensitive in relating cognitive performance to liability for psychosis (17,18). Using data from the Human Connectome Project for Early Psychosis (HCP-EP), we identified a relationship between DMPFC-somatomotor connectivity and ACPT performance that existed in individuals with early psychosis but not in controls. We then tested if this association was present even before the onset of psychosis using data from the North American Prodrome Longitudinal Study 2 (NAPLS2). We found that this relationship was only present in individuals at clinical high risk for psychosis (CHR-P) who would subsequently develop a psychotic disorder but not in those who did not develop psychosis (19). We therefore demonstrated a novel and reproducible relationship between DMPFC-somatomotor connectivity across multiple samples and multiple phases of illness. However, both HCP-EP and NAPLS2 were cross sectional studies limited to testing at a single timepoint.

To further test if DMPFC-somatomotor connectivity may be a promising treatment target for cognitive impairment, in the present study we tested if the relationship between DMPFC-somatomotor connectivity and ACPT performance would persist over time in a sample of individuals with early psychosis from HCP-EP and Neuroprogression datasets. The Neuroprogression dataset is a continuation of a subset of individuals from HCP-EP who were followed longitudinally over 4 years with repeat neuroimaging, clinical, and cognitive assessments. We hypothesized that DMPFC-somatomotor connectivity would be associated with ACPT performance across time. We also sought to investigate potential differences in the connectivity-cognition associations based on psychosis subtype (non-affective vs affective).

## Methods

### Participants

Data from 251 people with early psychosis and 90 matched healthy controls were enrolled. Individuals in the early psychosis group had a DSM-V diagnosis of a psychotic disorder with onset within the past 5 years prior to study entry. Eligible diagnoses in the non-affective psychosis cohort included schizophrenia, schizophreniform, schizoaffective, psychosis not otherwise specified, delusional disorder, or brief psychotic disorder, and diagnoses in the affective psychosis cohort included major depression with psychosis (single and recurrent episodes) or bipolar disorder with psychosis (including most recent episode depressed and manic types). Healthy controls had no lifetime or family history of psychosis or any other psychiatric disorder other than a history of anxiety and were not taking any psychiatric medication at the time of study entry (See Supplement for detailed Inclusion and Exclusion Criteria). All participants underwent baseline assessments of cognitive performance and neuroimaging, and a subset were followed longitudinally with repeated cognitive assessment and neuroimaging. Participants were assessed up to four times over 2 to 4 years. Prior to participation, all participants provided written informed consent in accordance with the institutional review boards of Indiana University, Indianapolis, Indiana and Partners Institutional Review Board Committee (now MGB, which served as the single IRB of record for Boston sites). Baseline data were collected across Boston-area and Indianapolis sites; follow-up data were collected at the Boston sites only.

### Cognitive Performance

The ACPT was used to assess cognitive performance at each visit (Figure 1) (20,21). The ACPT is comprised of 4 task conditions which differ based on degree of working memory and interference load. For each task condition, letters of the alphabet are presented monaurally at a rate of one letter per second for four blocks of 90 seconds. Subjects are required to respond to all target stimuli by lifting their index finger. ACPT total score was calculated by summing the vigilance, memory, and interference subscores, as previously (16). Additional methodological details are presented in the supplement.

**Figure 1.**
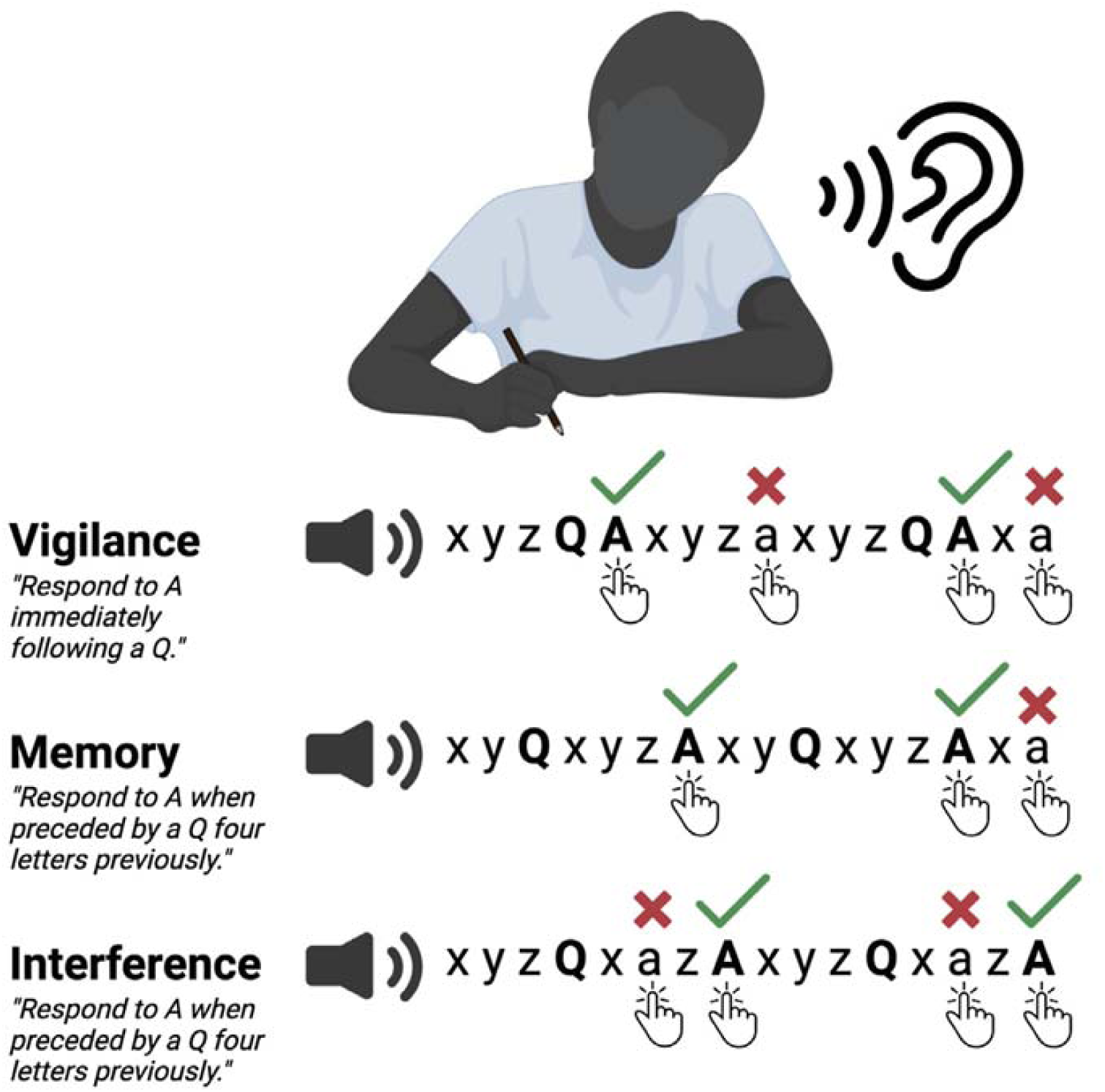
The Seidman Auditory Continuous Performance Task (ACPT). In the ACPT, individuals are presented with an auditory sensory stimulus (letters). There is a target response signal (the letter “A”) and a warning/cue signal (the letter “Q”). The ACPT contains several conditions that differ based on their degree of working memory and interference load. Working memory load is defined as the number of letters between the warning/cue and the target. To make the task more difficult, competing information (i.e., “interference”) is added to increase task demands within a continuous cognitive updating (i.e., CPT) framework. Interference load i defined by the number of distracters (Q’s and A’s) embedded between the cue and the target (18). Additional methodological details are presented in the Supplement. We used the ACPT total score as a summary measure of cognitive performance. The ACPT total score was calculated by summing the vigilance, memory, and interference sub scores.

The NIH Toolbox Cognition battery (22) was administered to assess multiple domains of cognitive functioning. The NIH Toolbox Cognition battery yields individual subtests scores, an overall composite, and fluid and crystallized composite scores. The fluid cognition composite (used in sensitivity analyses) reflects cognitive skills required to solve problems in real-time (e.g. information processing speed; working memory), whereas crystallized cognition reflects the ability to tap into knowledge learned previously (e.g. defining vocabulary words). The fluid composite score includes averaged scaled scores from the following subtests: Dimensional Change Card Sort, Flanker Inhibitory Control and Attention, Picture Sequence Memory, List Sorting Working Memory, and Pattern Comparison Processing Speed tests. Together these tasks measure executive functioning, inhibitory control, attention, episodic memory, working memory, and processing speed.

### Psychosis Symptoms

Psychosis symptoms were measured using the Positive and Negative Syndrome Scale (PANSS) (23). The Marder factor analysis was used to calculate positive, negative, and general psychopathology subscores (24).

### Antipsychotic Medication Dose

Participants self-reported their antipsychotic medication dose, which was converted to a chlorpromazine equivalent dose (CPZeq) using the Gardner approach (25).

### MRI Acquisition

Imaging was conducted on Siemens 3.0-T MRI or PRISMA 3T systems (Munich, Germany). Briefly, 0.8-mm^3^ T1-weighted anatomical scans were acquired, and resting-state functional runs of approximately 6 minutes were acquired from all participants (420 time points, 0.8-second repetition time, 2-mm^3^ voxels). Details in supplement.

### MRI Data Processing

All analyses were preprocessed using the Data Processing and Analysis for Brain Imaging toolbox ((26); http://rfmri.org/dpabi). As a quality control metric, resting-state scans that exceeded motion thresholds (>3mm mean translation or >3 degrees mean rotation) were discarded. Individual time points with framewise displacement >0.2mm were discarded, and scans with >50% of volumes removed for framewise displacement were discarded. All data were preprocessed to remove motion (24-parameter), CSF signals, white matter signals, and an overall linear trend. A bandpass filter was applied (0.01-0.08 Hz). Data were normalized using the DARTEL toolbox (http://www.neurometrika.org/node/34) into Montreal Neurological Institute (MNI) space and smoothed with an 8-mm full-width half-maximum kernel. Voxels within a pre-defined (MNI) gray matter mask were used for further analysis. Data were resampled into 4mm isotropic resolution. Details in supplement.

### MRI Analysis

#### Seed-Based Connectivity Analysis

To calculate DMPFC-somatomotor connectivity, we measured the BOLD correlation between the previously identified prefrontal region and a 6mm sphere (seed) placed at the previously identified location of maximal connectivity-cognition association in the somatomotor cortex (MNI X= +4, Y = −40, Z = +68). We then modeled the association between DMPFC-somatomotor connectivity with ACPT total score. Details in supplement.

### Statistical Approach

To compare continuous outcomes based on dichotomous variables, t-tests were used. Pearson’s chi squared test was used to compare categorical outcomes based on categorical variables. Linear mixed models were used to determine the relationships between functional connectivity and cognitive performance. ACPT performance was modeled as the outcome including DMPFC-somatomotor connectivity, time from baseline, and antipsychotic medication dose (chlorpromazine equivalents) as covariates and a random intercept for subject. To test if psychosis subtype (affective vs. nonaffective) affected the relationship between DMPFC-somatomotor connectivity and ACPT performance, we ran separate models testing if the connectivity by psychosis subtype interaction predicted ACPT performance. To test the specificity of the relationship between DMPFC-somatomotor connectivity and ACPT performance, we also modeled DMPFC-connectivity based on ACPT performance and an alternative measure of cognitive performance (NIH toolbox fluid cognition). Tests were performed using Kenward-Rogers adjustment for the degrees of freedom. Data were assumed to be missing at random. All analyses were conducted in RStudio (Version 2025-06-13 ucrt).

## Results

After quality control, a total of 325 participants contributed functional MRI and ACPT data for analysis. There were 241 participants in the early psychosis group and 84 control participants. A total of 22 individuals provided longitudinal data for analysis (i.e., multiple timepoints of cognitive assessment and neuroimaging). There were no significant differences in age or sex between participants with psychosis and control participants (Table 1).

**Table 1.** Sample Demographics. Individuals with early psychosis (n=251) and matched healthy controls (90) were enrolled at baseline and underwent neuroimaging and cognitive assessment. ACPT: Auditory Continuous Performance Task; PANSS: Positive and Negative Syndrome Scale.

| **Variable** | **Psychosis (n=251)** | **Control (n=90)** | ***p*** |
| --- | --- | --- | --- |
| Age, M (SD) | 23.9 (3.8) | 23.6 (3.5) | 0.70 |
| Sex, n Female (%) | 63 (50) | 25 (64.1) | 0.18 |
| PANSS Total, M (SD) | 44.6 (10.9) | -- | -- |
| Taking Antipsychotic Medication (%) | 68.3% | 0% | -- |
| ACPT Total, M (SD) | 259.2 (33.5) | 277.8 (16.5) | <0.001 |

### ACPT Performance Changes Over Time

At baseline, the psychosis group had worse ACPT total performance than controls (t(262.61)=7.58, p<.001). Within the psychosis group, individuals with nonaffective psychosis performed worse on the ACPT (t(216.25)=-5.89, p<.001).

In a model predicting ACPT total performance based on time and diagnosis (psychosis vs. control), time (Est=0.021, SE=0.0051, t(277.40)=4.09, p<.0001) and psychosis diagnosis (Est=- 24.96, SE=4.29, t(307.39)=-5.81, p<.0001) were significant predictors (Figure 2A, Supplemental Table 1). When the timepoint by diagnosis interaction was included in the model, it was not significant (p>.05, Supplemental Table 2).

**Figure 2.**
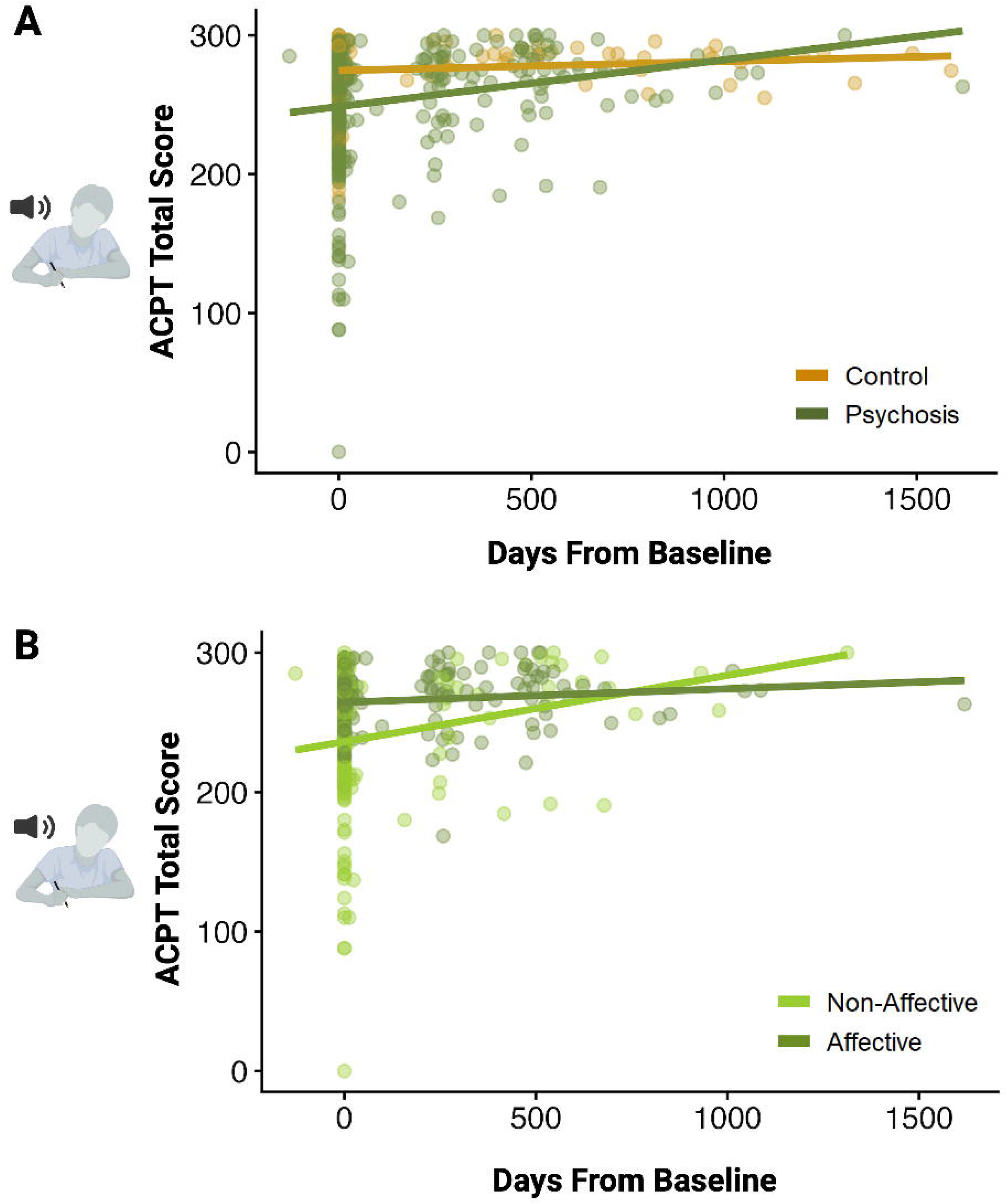
Individuals with Psychosis Perform Worse on the ACPT than Controls, but Performance Improves Over Time. At baseline, the psychosis group performs worse on the ACPT compared to controls (247.2 vs 273.7, p<.001). In a model predicting ACPT total performance based on time and diagnosis (psychosis vs. control), time (p<.0001) and psychosis diagnosis (p<.0001) were significant predictors (Figure 2A). In the psychosis sample, time (p=.0037) and affective psychosis diagnosis (p<.0001) predicted better performance (Figure 2B).

In the psychosis sample, we predicted ACPT total performance based on time and psychosis subtype (affective or nonaffective psychosis). In this model, time (Est=0.020, SE=0.0068, t(173.80)=2.94, p=.0037) and affective psychosis diagnosis (Est=22.55, SE=4.70, t(215.70)=4.80, p<.0001) predicted better performance (Figure 2B, Supplemental Table 3). When the timepoint by psychosis subtype interaction was included in the model, it was not significant (p>.05, Supplemental Table 4).

### Across Time, DMPFC-Somatomotor Connectivity Predicts ACPT Performance

Because of the variability in timing of assessments, we calculated time from baseline as a continuous variable. We first modeled ACPT performance based on DMPFC-somatomotor connectivity, psychosis subtype (non-affective vs affective), and days from baseline, while controlling for antipsychotic medication dose (CPZeq) and including a random effect for subject. Affective psychosis (Est=28.82, SE=8.33, t(137.76)=3.46, p<.001), time from baseline (Est=0.021, SE=0.0070, t(35.47)=3.02, p=.0047), and antipsychotic medication dose (CPZeq, Est=-0.032, SE=0.013, t(87.59)=-2.42, p=.017) were significant predictors of ACPT performance (Figure 3, Supplemental Table 5). In this model, DMPFC-somatomotor connectivity was not a significant predictor (p=.21).

**Figure 3.**
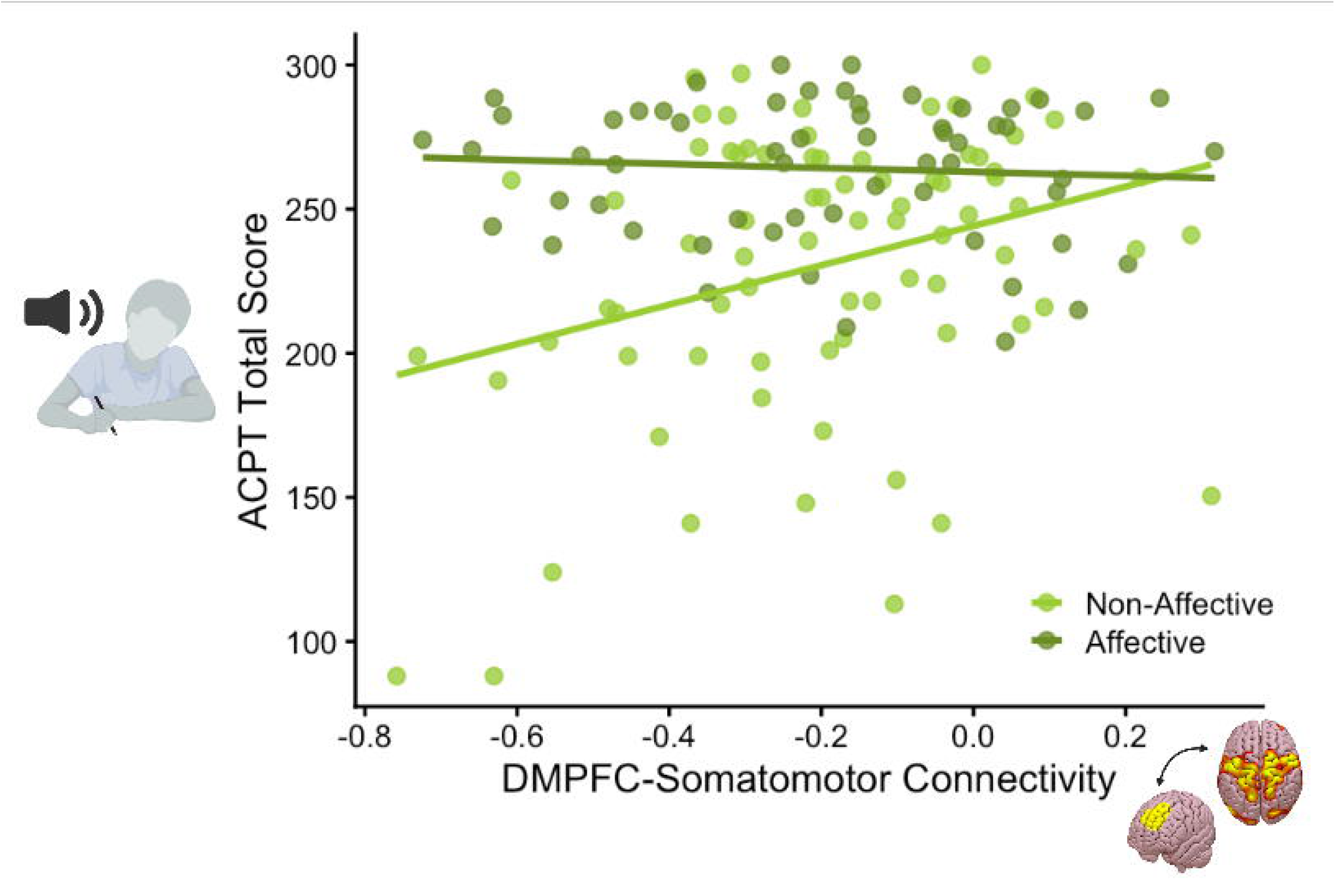
The Relationship Between DMPFC-Somatomotor Connectivity and ACPT Performance is Specific to Non-Affective Psychosis. In this sample of individuals with early psychosis and longitudinal neurocognitive assessment and resting-state neuroimaging, we calculated DMPFC-somatomotor connectivity using our previously identified DMPFC cluster and somatomotor cortex seed. Because of the variability in timing of assessments, we calculated time from baseline as a continuous variable. In a model predicting ACPT performance, we observed a significant interaction effect of DMPFC-somatomotor connectivity*psychosis subtype (p=.0079) such that DMPFC-somatomotor connectivity predicted ACPT performance only in individuals with non-affective psychosis (p=.0051), not in affective psychosis (p>.05).

However, when we tested if the relationship between DMPFC-somatomotor connectivity and ACPT performance was affected by psychosis subtype, we observed a significant interaction effect (Est=-53.36, SE=19.48, t(67.80)=-2.74, p=.0079, Supplemental Table 6). In posthoc analyses, there was a significant relationship between DMPFC-somatomotor connectivity and ACPT performance only for individuals with non-affective psychosis (Est=47.67, SE=16.7, t(120.5)=2.86, p=.0051, Figure 3, Supplemental Table 7).

### ACPT, not Fluid Cognition, Predicts DMPFC-Somatomotor Connectivity

We then sought to test if the association between ACPT performance and DMPFC-somatomotor connectivity was specific to the ACPT, or if DMPFC-somatomotor connectivity might also predict another measure of cognitive performance (fluid cognition). To do this, we predicted DMPFC-somatomotor connectivity based on fluid cognition, ACPT performance, psychosis subtype (non-affective vs. affective), antipsychotic medication dose (CPZeq), and time and including a random effect for subject. In this model, only ACPT performance was a significant predictor of connectivity (Est=0.0015, SE=0.00060, t(127.90)=2.41, p=.017, Figure 4, Supplemental Table 8). When we included the interaction between ACPT performance and psychosis subtype, it did not significantly predict DMPFC-somatomotor connectivity (p>.05, Supplemental Table 9). This specificity analysis suggests the relationship between DMPFC-somatomotor connectivity and ACPT performance is in fact specific to the ACPT rather than a more generic measure of cognitive performance.

**Figure 4.**
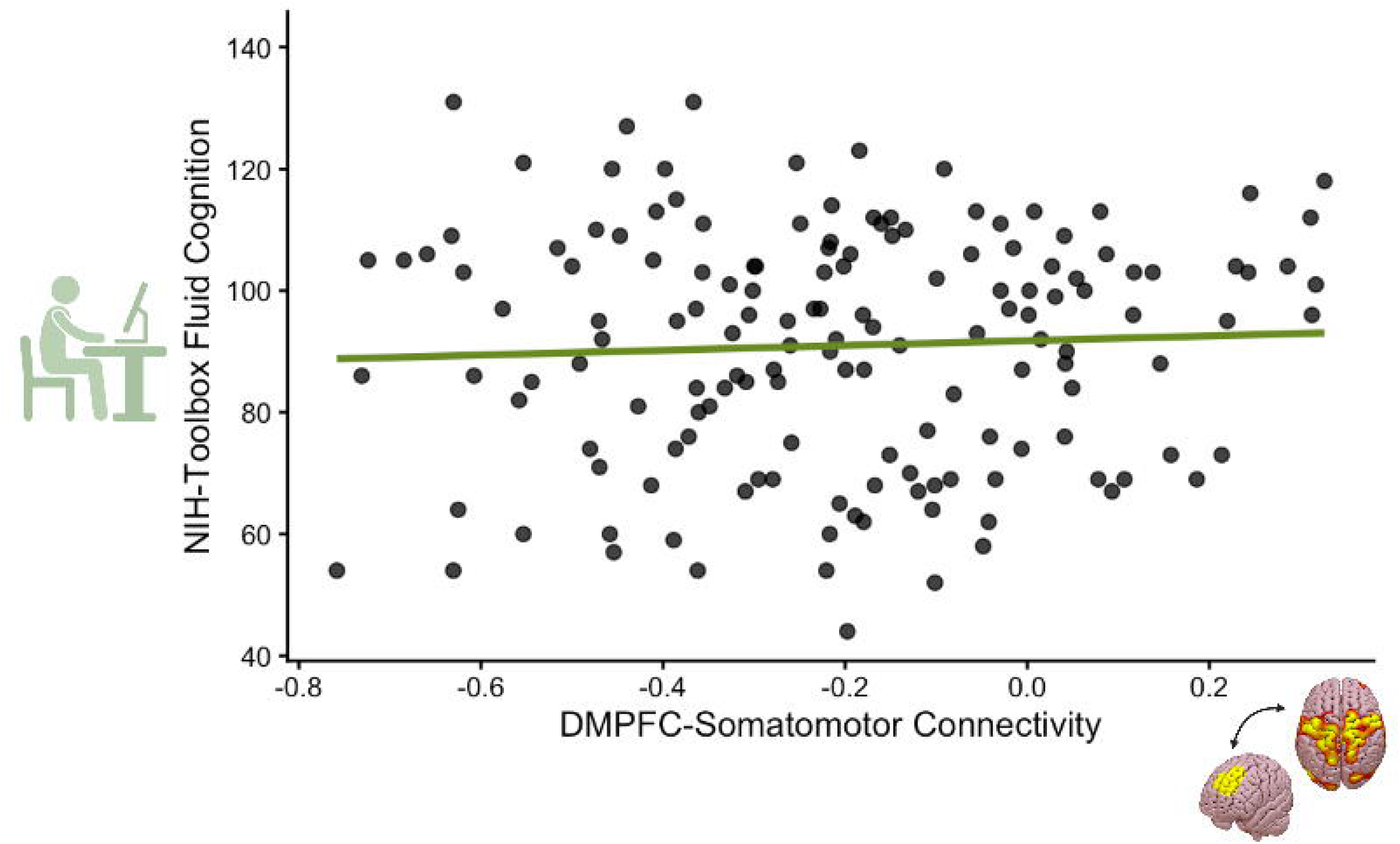
General Cognitive Performance Does Not Predict DMPFC-Somatomotor Connectivity. We tested if the association between ACPT performance and DMPFC-somatomotor connectivity was specific to the ACPT, or if DMPFC-somatomotor connectivity might also predict another measure of cognitive performance (fluid cognition). To do this, we predicted DMPFC-somatomotor connectivity based on fluid cognition, ACPT performance, psychosis subtype (non-affective vs. affective), antipsychotic medication dose (CPZeq), and time and including a random effect for subject. In this model, only ACPT performance was a significant predictor of connectivity (p=.017), not fluid cognition (p>.05). This specificity analysis suggests the relationship between DMPFC-somatomotor connectivity and ACPT performance is in fact specific to the ACPT rather than a more generic measure of cognitive performance.

## Discussion

Cognitive impairment is a leading cause of disability and functional impairment in people with psychosis but has limited effective treatments. Neuroimaging studies in psychosis have struggled to identify reliable neural markers of cognitive impairment that could serve as potential treatment targets. A promising neural marker would need to be readily and reproducibly identifiable, exist across a population of people with psychosis, and be reliably linked to cognitive deficits within individuals across time. A neural marker meeting these criteria has remained elusive until now.

In previous work, we identified a reproducible association between cognitive performance and DMPFC-somatomotor connectivity in two independent samples of individuals across the psychosis spectrum (19). Now, in this prospective cohort study, we demonstrated that the relationship between ACPT performance and DMPFC-somatomotor connectivity remains stable over time, where better performance is associated with higher DMPFC-somatomotor connectivity. Prior to the present study, previous work observed this relationship in *cross-sectional* datasets of individual with psychosis (19). Here, we expanded this work to demonstrate that increased prefrontal somatomotor activity is associated with better cognitive performance *longitudinally* in an early psychosis sample. Importantly, DMPFC-somatomotor connectivity did not predict fluid cognition, a more general measure of cognitive performance. This reaffirms the specificity of the relationship between DMPFC-somatomotor connectivity and performance on this particular cognitive task, the ACPT, which was specifically developed to assess cognitive deficits in psychosis (21,27,28).

The results of this study suggest that DMPFC-somatomotor connectivity is a stable neural marker of cognitive deficits in schizophrenia. We have now shown in multiple studies that the relationship between DMPFC-somatomotor connectivity and ACPT performance is specific to psychotic disorders (i.e., does not exist in healthy controls), reliable over time, and reproducible across multiple samples.

Attention is among the most central cognitive deficits observed in schizophrenia and one of the most extensively studied (29). Impaired auditory attention is a core deficit present in schizophrenia, their non-psychotic relatives, and individuals at risk for psychosis (21,27,28). People with schizophrenia have well-established deficits in auditory processing, including dysfunction in auditory processing areas of the temporal lobe (30) and abnormal auditory event related potentials (ERPs) (31). Auditory stimuli used in experiments testing prepulse inhibition (32), P50 (33), and mismatch negativity (34) yield abnormal ERPs in schizophrenia and their first-degree relatives. Sustained attention has been measured using various forms of continuous performance tasks (35–39). Given the established deficits in attentional performance and auditory processing, the ACPT was specifically developed to assess sustained auditory attention with an increasing working memory load in individuals with or at risk for schizophrenia. Therefore, the ACPT detects pathognomonic cognitive deficits in schizophrenia and may enable reliable identification of novel circuits linked to cognitive deficits in schizophrenia.

Though we have used resting-state neuroimaging, the ACPT has been performed in-scanner using task-based fMRI in healthy and psychosis populations. Multiple studies have observed DMPFC (20,40) and somatomotor network activation (20,40,41) during ACPT performance, consistent with our identified DMPFC and somatomotor regions. This suggests the DMPFC and somatomotor network are related to cognition during task and rest.

The DMPFC is critically involved in cognition and converging evidence suggests the DMPFC plays a central role in sustaining attention and exerting cognitive control when behavior needs to be regulated over a temporal delay. Specifically, neuroimaging studies have linked the DMPFC to expectation and anticipatory attention, supporting an alert state in preparation for an upcoming stimulus (42). Other work has shown that the DMPFC is engaged when actions must be selected according to task-relevant stimuli and contextual demands, including sensory-motor control (43). Consistent with this, rodent studies have demonstrated that the DMPFC supports response inhibition during tasks requiring subjects to wait before responding, through top-down modulation of motor cortex activity during delay periods (44). From a network perspective, the DMPFC serves as an important hub that links the frontoparietal and cingulo-opercular networks (43), networks critical for top-down cognitive control and tonic attention, respectively (45,46). Together, these findings suggest that the DMPFC supports the sustained attentional engagement, response preparation, and inhibitory control necessary for maintaining task performance across delays through engagement with somatomotor regions – processes that are important for successful performance on the ACPT. Consequently, disrupted DMPFC function or connectivity may contribute to cognitive impairments observed in people with schizophrenia. Indeed, abnormal activity in the DMPFC has been associated with cognitive deficits in individuals with schizophrenia (47) and suggests stimulating the DMPFC may enhance attentional control via its connections to somatomotor network (43).

When we investigated differences in affective versus non-affective diagnoses, higher DMPFC-somatomotor connectivity was associated with better performance in the non-affective group but not the affective group. It is unclear if this relationship is truly specific to non-affective psychosis, or if the relationship does generalize to individuals with psychosis but is more pronounced in individuals with greater cognitive impairment. This should be explored further in larger, longitudinal studies and interventional clinical trials that modulate DMPFC-somatomotor connectivity in a broad psychosis sample.

As expected, the psychosis group performed significantly worse than the control group at all time points on the ACPT (Figure 2). This finding aligns with previous literature demonstrating impaired attentional control in psychosis populations (19,48,49). Within the psychosis group, affective and non-affective psychoses were associated with differences in performance, where participants with affective psychosis performed better than participants with non-affective psychosis. This is consistent with previous literature identifying more pronounced cognitive deficits in people with non-affective psychosis compared to affective psychosis (19,50,51). Research has previously suggested that individuals with affective psychosis may be “cognitively spared” from the core cognitive deficits associated with non-affective diagnoses such as schizophrenia and related disorders (51). In affective psychosis, psychotic symptoms often occur alongside mood disorders and cognitive impairment is not always a core deficit observed in this population (50,52). It is possible that the underlying pathophysiology and extent of cognitive impairment between affective and non-affective psychosis may differ, leading to differences in attentional control performance.

It is important to acknowledge the limitations of the present study. The individuals with psychosis included in our analysis are a relatively stable population with an average PANSS score of 48.8 across all visits. Since the participants in this study were recruited within their first five years of illness, they may be less symptomatic than chronic populations (53). Future studies examining this relationship across all stages of illness are needed to confirm that this finding extends beyond early psychosis.

There are many unique strengths of this longitudinal, multi-site analysis. The pairing of the ACPT, a sophisticated, well-validated task, with repeated neuroimaging allows for a unique examination of the stability of brain-behavior relationships as individuals progress through the early stages of illness. Previous longitudinal studies have shown that structural and functional brain changes occur most rapidly during early psychosis, particularly within prefrontal regions, and that large-scale network abnormalities are already present at illness onset (54–56). The persistent association between DMPFC-somatomotor connectivity and ACPT performance across repeated assessments suggests that this brain-behavior relationship remains stable and reproducible despite the substantial neurobiological change occurring during early psychosis. Future studies should determine whether this association extends into the chronic stages of illness.

The results of this study suggest that DMPFC-somatomotor connectivity is a stable neural marker of cognitive deficits in schizophrenia. We have now shown in multiple studies that the relationship between DMPFC-somatomotor connectivity and ACPT performance is specific to psychotic disorders, reliable over time, and reproducible across multiple samples. A critical next step is to determine whether this marker can be directly modulated, which would establish a novel, mechanistically informed treatment for cognitive impairment in psychosis.

## Data Availability

Data are available on the NIMH National Data Archive and on reasonable request to the authors.

## Acknowledgments

This work was supported by National Institutes of Health (NIH) grants R01 MH116170 to Dr. Brady, R01 MH117012 to Dr. Lewandowski, and K23DA059690 to Dr. Ward.

## References

1. Kuperberg G, Heckers S. Schizophrenia and cognitive function. Curr Opin Neurobiol. 2000 Apr 1;10(2):205–10. doi:10.1016/S0959-4388(00)00068-4

2. Bouwmans C, de Sonneville C, Mulder CL, Hakkaart-van Roijen L. Employment and the associated impact on quality of life in people diagnosed with schizophrenia. Neuropsychiatr Dis Treat. 2015 Aug 18;11:2125–42. doi:10.2147/NDT.S83546 PubMed PMID: 26316759; PubMed Central PMCID: PMC4547637.

3. Dickson H, Hedges EP, Ma SY, Cullen AE, MacCabe JH, Kempton MJ, et al. Academic achievement and schizophrenia: a systematic meta-analysis. Psychol Med. 2020 Sep;50(12):1949–65. doi:10.1017/S0033291720002354 PubMed PMID: 32684198.

4. Abplanalp SJ, Catalano LT, Green MF. Advancing the Measurement of Social Functioning in Schizophrenia: Applications of Egocentric Social Network Analysis. Schizophr Bull. 2024 Jul 27;50(4):723–30. doi:10.1093/schbul/sbae082 PubMed PMID: 38828486; PubMed Central PMCID: PMC11283182.

5. Lencz T, Smith CW, McLaughlin D, Auther A, Nakayama E, Hovey L, et al. Generalized and specific neurocognitive deficits in prodromal schizophrenia. Biol Psychiatry. 2006 May 1;59(9):863–71. doi:10.1016/j.biopsych.2005.09.005 PubMed PMID: 16325151.

6. Fatouros-Bergman H, Cervenka S, Flyckt L, Edman G, Farde L. Meta-analysis of cognitive performance in drug-naïve patients with schizophrenia. Schizophr Res. 2014 Sep 1;158(1):156–62. doi:10.1016/j.schres.2014.06.034

7. Li J, Lu X, Du S, Hu W, Xiao D, He B, et al. Effects of varied rTMS frequencies on cognitive function in individuals with chronic schizophrenia: A double-blind randomized controlled trial. J Psychiatr Res. 2025 Sep 1;189:33–41. doi:10.1016/j.jpsychires.2025.05.052

8. Xiu MH, Guan HY, Zhao JM, Wang KQ, Pan YF, Su XR, et al. Cognitive Enhancing Effect of High-Frequency Neuronavigated rTMS in Chronic Schizophrenia Patients With Predominant Negative Symptoms: A Double-Blind Controlled 32-Week Follow-up Study. Schizophr Bull. 2020 Sep 21;46(5):1219–30. doi:10.1093/schbul/sbaa035

9. Chechko N, Cieslik EC, Müller VI, Nickl-Jockschat T, Derntl B, Kogler L, et al. Differential Resting-State Connectivity Patterns of the Right Anterior and Posterior Dorsolateral Prefrontal Cortices (DLPFC) in Schizophrenia. Front Psychiatry. 2018 May 28;9. doi:10.3389/fpsyt.2018.00211

10. Birchwood M, Todd P, Jackson C. Early intervention in psychosis: The critical period hypothesis. Br J Psychiatry. 1998 Jun;172(S33):53–9. doi:10.1192/S0007125000297663

11. Pardo-de-Santayana G, Vázquez-Bourgon J, Gómez-Revuelta M, Ayesa-Arriola R, Ortiz-Garcia de la Foz V, Crespo-Facorro B, et al. Duration of active psychosis during early phases of the illness and functional outcome: The PAFIP 10-year follow-up study. Schizophr Res. 2020 Jun 1;220:240–7. doi:10.1016/j.schres.2020.03.009

12. Keefe RSE. Why are there no approved treatments for cognitive impairment in schizophrenia? World Psychiatry. 2019 Jun;18(2):167–8. doi:10.1002/wps.20648 PubMed PMID: 31059617; PubMed Central PMCID: PMC6502426.

13. Mana L, Schwartz-Pallejà M, Vila-Vidal M, Deco G. Overview on cognitive impairment in psychotic disorders: From impaired microcircuits to dysconnectivity. Schizophr Res. 2024 Jul 1;269:132–43. doi:10.1016/j.schres.2024.05.008

14. Altered prefrontal activity and connectivity predict different cognitive deficits in schizophrenia [Internet]. doi:10.1002/hbm.22935

15. Marek S, Tervo-Clemmens B, Calabro FJ, Montez DF, Kay BP, Hatoum AS, et al. Reproducible brain-wide association studies require thousands of individuals. Nature. 2022 Mar;603(7902):654–60. doi:10.1038/s41586-022-04492-9 PubMed PMID: 35296861; PubMed Central PMCID: PMC8991999.

16. Ward HB, Beermann A, Xie J, Yildiz G, Felix KM, Addington J, et al. Robust Brain Correlates of Cognitive Performance in Psychosis and Its Prodrome. Biol Psychiatry. 2025 Jan 15;Brain Structural and Functional Connectivity in Psychosis: Relationship to Clinical Outcomes97(2):139–47. doi:10.1016/j.biopsych.2024.07.012

17. Neff MA, Raffensperger KV, Moore RC, Depp CA, Ackerman RA, Pinkham AE, et al. Predictions of different elements of everyday functional outcomes in bipolar disorder and schizophrenia: Cognition, social cognition, clinical symptoms, and mood states as predictors. Schizophr Res. 2026 Feb 1;288:77–85. doi:10.1016/j.schres.2025.12.018

18. Seidman LJ, Meyer EC, Giuliano AJ, Breiter HC, Goldstein JM, Kremen WS, et al. Auditory working memory impairments in individuals at familial high risk for schizophrenia. Neuropsychology. 2012;26(3):288–303. doi:10.1037/a0027970

19. Ward HB, Beermann A, Xie J, Yildiz G, Felix KM, Addington J, et al. Robust Brain Correlates of Cognitive Performance in Psychosis and Its Prodrome. Biol Psychiatry. 2025 Jan 15;97(2):139–47. doi:10.1016/j.biopsych.2024.07.012 PubMed PMID: 39032726.

20. Seidman LJ, Breiter HC, Goodman JM, Goldstein JM, Woodruff PW, O’Craven K, et al. A functional magnetic resonance imaging study of auditory vigilance with low and high information processing demands. Neuropsychology. 1998 Oct;12(4):505–18. doi:10.1037//0894-4105.12.4.505 PubMed PMID: 9805320.

21. Seidman LJ, Meyer EC, Giuliano AJ, Breiter HC, Goldstein JM, Kremen WS, et al. Auditory working memory impairments in individuals at familial high risk for schizophrenia. Neuropsychology. 2012 May;26(3):288–303. doi:10.1037/a0027970 PubMed PMID: 22563872; PubMed Central PMCID: PMC3539430.

22. Nuechterlein KH, Green MF, Kern RS, Baade LE, Barch DM, Cohen JD, et al. The MATRICS Consensus Cognitive Battery, part 1: test selection, reliability, and validity. Am J Psychiatry. 2008 Feb;165(2):203–13. doi:10.1176/appi.ajp.2007.07010042 PubMed PMID: 18172019.

23. Kay SR, Fiszbein A, Opler LA. The positive and negative syndrome scale (PANSS) for schizophrenia. Schizophr Bull. 1987;13(2):261–76. doi:10.1093/schbul/13.2.261 PubMed PMID: 3616518.

24. Marder SR, Davis JM, Chouinard G. The effects of risperidone on the five dimensions of schizophrenia derived by factor analysis: combined results of the North American trials. J Clin Psychiatry. 1997 Dec;58(12):538–46. doi:10.4088/jcp.v58n1205 PubMed PMID: 9448657.

25. Gardner DM, Murphy AL, O’Donnell H, Centorrino F, Baldessarini RJ. International consensus study of antipsychotic dosing. Am J Psychiatry. 2010 Jun;167(6):686–93. doi:10.1176/appi.ajp.2009.09060802 PubMed PMID: 20360319.

26. Yan CG, Wang XD, Zuo XN, Zang YF. DPABI: Data Processing & Analysis for (Resting-State) Brain Imaging. Neuroinformatics. 2016 Jul 1;14(3):339–51. doi:10.1007/s12021-016-9299-4

27. Faraone SV, Seidman LJ, Kremen WS, Pepple JR, Lyons MJ, Tsuang MT. Neuropsychological functioning among the nonpsychotic relatives of schizophrenic patients: a diagnostic efficiency analysis. J Abnorm Psychol. 1995 May;104(2):286–304. doi:10.1037//0021-843x.104.2.286 PubMed PMID: 7790631.

28. Seidman LJ, Shapiro DI, Stone WS, Woodberry KA, Ronzio A, Cornblatt BA, et al. Association of Neurocognition With Transition to Psychosis: Baseline Functioning in the Second Phase of the North American Prodrome Longitudinal Study. JAMA Psychiatry. 2016 Dec 1;73(12):1239–48. doi:10.1001/jamapsychiatry.2016.2479 PubMed PMID: 27806157; PubMed Central PMCID: PMC5511703.

29. Neuropsychological and Structural Neuroimaging Endophenotypes in Schizophrenia – Stone - Major Reference Works - Wiley Online Library [Internet]. [cited 2026 Aug 21]. Available from: https://onlinelibrary.wiley.com/doi/10.1002/9781119125556.devpsy224

30. Hirayasu Y, McCarley RW, Salisbury DF, Tanaka S, Kwon JS, Frumin M, et al. Planum temporale and Heschl gyrus volume reduction in schizophrenia: a magnetic resonance imaging study of first-episode patients. Arch Gen Psychiatry. 2000 Jul;57(7):692–9. doi:10.1001/archpsyc.57.7.692 PubMed PMID: 10891040; PubMed Central PMCID: PMC2850271.

31. Meta analysis of P300 and schizophrenia: Patients, paradigms, and practical implications - Jeon - 2003 - Psychophysiology - Wiley Online Library [Internet]. [cited 2026 Aug 21]. Available from: https://onlinelibrary.wiley.com/doi/10.1111/1469-8986.00070

32. Braff D, Stone C, Callaway E, Geyer M, Glick I, Bali L. Prestimulus effects on human startle reflex in normals and schizophrenics. Psychophysiology. 1978 Jul;15(4):339–43. doi:10.1111/j.1469-8986.1978.tb01390.x PubMed PMID: 693742.

33. Adler LE, Pachtman E, Franks RD, Pecevich M, Waldo MC, Freedman R. Neurophysiological evidence for a defect in neuronal mechanisms involved in sensory gating in schizophrenia. Biol Psychiatry. 1982 Jun;17(6):639–54. PubMed PMID: 7104417.

34. Michie PT. What has MMN revealed about the auditory system in schizophrenia? Int J Psychophysiol Off J Int Organ Psychophysiol. 2001 Oct;42(2):177–94. doi:10.1016/s0167-8760(01)00166-0 PubMed PMID: 11587775.

35. Nuechterlein KH, Green MF, Kern RS, Baade LE, Barch DM, Cohen JD, et al. The MATRICS Consensus Cognitive Battery, Part 1: Test Selection, Reliability, and Validity. Am J Psychiatry. 2008 Feb;165(2):203–13. doi:10.1176/appi.ajp.2007.07010042

36. Cornblatt BA, Keilp JG. Impaired attention, genetics, and the pathophysiology of schizophrenia. Schizophr Bull. 1994;20(1):31–46. doi:10.1093/schbul/20.1.31 PubMed PMID: 8197420.

37. Gur RC, Braff DL, Calkins ME, Dobie DJ, Freedman R, Green MF, et al. Neurocognitive performance in family-based and case-control studies of schizophrenia. Schizophr Res. 2015 Apr;163(1–3):17–23. doi:10.1016/j.schres.2014.10.049 PubMed PMID: 25432636; PubMed Central PMCID: PMC4441547.

38. Nuechterlein KH, Parasuraman R, Jiang Q. Visual Sustained Attention: Image Degradation Produces Rapid Sensitivity Decrement Over Time. Science. 1983 Apr 15;220(4594):327–9. doi:10.1126/science.6836276

39. López-Luengo B, González-Andrade A, García-Cobo M. Not All Differences between Patients with Schizophrenia and Healthy Subjects Are Pathological: Performance on the Conners’ Continuous Performance Test. Arch Clin Neuropsychol. 2016 Dec 24;31(8):983–95. doi:10.1093/arclin/acw075

40. Distinct cortical networks activated by auditory attention and working memory load - ClinicalKey [Internet]. [cited 2026 Aug 21]. Available from: https://www.clinicalkey.com/#!/content/playContent/1-s2.0-S1053811913008434?returnurl= https://linkinghub.elsevier.com%2Fretrieve%2Fpii%2FS1053811913008434%3Fshowall%3Dtrue&referrer=

41. Goldstein JM, Jerram M, Poldrack R, Anagnoson R, Breiter HC, Makris N, et al. Sex differences in prefrontal cortical brain activity during fMRI of auditory verbal working memory. Neuropsychology. 2005 Jul;19(4):509–19. doi:10.1037/0894-4105.19.4.509 PubMed PMID: 16060826.

42. Walter M, Matthiä C, Wiebking C, Rotte M, Tempelmann C, Bogerts B, et al. Preceding attention and the dorsomedial prefrontal cortex: process specificity versus domain dependence. Hum Brain Mapp. 2009 Jan;30(1):312–26. doi:10.1002/hbm.20506 PubMed PMID: 18072281; PubMed Central PMCID: PMC6870847.

43. Wood JL, Nee DE. Cingulo-Opercular Subnetworks Motivate Frontoparietal Subnetworks during Distinct Cognitive Control Demands. J Neurosci. 2023 Feb 15;43(7):1225–37. doi:10.1523/JNEUROSCI.1314-22.2022 PubMed PMID: 36609452.

44. Narayanan NS, Laubach M. Top-Down Control of Motor Cortex Ensembles by Dorsomedial Prefrontal Cortex. Neuron. 2006 Dec 7;52(5):921–31. doi:10.1016/j.neuron.2006.10.021 PubMed PMID: 17145511.

45. Sadaghiani S, D’Esposito M. Functional Characterization of the Cingulo-Opercular Network in the Maintenance of Tonic Alertness. Cereb Cortex. 2015 Sep 1;25(9):2763–73. doi:10.1093/cercor/bhu072

46. Marek S, Dosenbach NUF. The frontoparietal network: function, electrophysiology, and importance of individual precision mapping. Dialogues Clin Neurosci. 2018 Jun 30;20(2):133–40. doi:10.31887/DCNS.2018.20.2/smarek PubMed PMID: 30250390.

47. Krawitz A, Braver TS, Barch DM, Brown JW. Impaired error-likelihood prediction in medial prefrontal cortex in schizophrenia. NeuroImage. 2011 Jan 15;54(2):1506–17. doi:10.1016/j.neuroimage.2010.09.027

48. Ikuta T, Robinson DG, Gallego JA, Peters BD, Gruner P, Kane J, et al. Subcortical Modulation of Attentional Control by Second-Generation Antipsychotics in First-Episode Psychosis. Psychiatry Res. 2014 Feb 28;221(2):127–34. doi:10.1016/j.pscychresns.2013.09.010 PubMed PMID: 24120303; PubMed Central PMCID: PMC3946302.

49. Cornblatt BA, Erlenmeyer-Kimling L. Global attentional deviance as a marker of risk for schizophrenia: Specificity and predictive validity. J Abnorm Psychol. 1985;94(4):470–86. doi:10.1037/0021-843X.94.4.470

50. Bora E, Yucel M, Pantelis C. Cognitive functioning in schizophrenia, schizoaffective disorder and affective psychoses: meta-analytic study. Br J Psychiatry J Ment Sci. 2009 Dec;195(6):475–82. doi:10.1192/bjp.bp.108.055731 PubMed PMID: 19949193.

51. Bracher KM, Wohlschlaeger A, Koch K, Knolle F. Cognitive subgroups of affective and non-affective psychosis show differences in medication and cortico-subcortical brain networks. Sci Rep. 2024 Sep 2;14(1):20314. doi:10.1038/s41598-024-71316-3

52. Bora E, Yücel M, Pantelis C. Cognitive impairment in schizophrenia and affective psychoses: implications for DSM-V criteria and beyond. Schizophr Bull. 2010 Jan;36(1):36–42. doi:10.1093/schbul/sbp094 PubMed PMID: 19776206; PubMed Central PMCID: PMC2800141.

53. Tang B, Yao L, Strawn JR, Zhang W, Lui S. Neurostructural, Neurofunctional, and Clinical Features of Chronic, Untreated Schizophrenia: A Narrative Review. Schizophr Bull. 2025 Mar 14;51(2):366–78. doi:10.1093/schbul/sbae152 PubMed PMID: 39212651; PubMed Central PMCID: PMC11908860.

54. Andreasen NC, Nopoulos P, Magnotta V, Pierson R, Ziebell S, Ho BC. Progressive brain change in schizophrenia: a prospective longitudinal study of first-episode schizophrenia. Biol Psychiatry. 2011 Oct 1;70(7):672–9. doi:10.1016/j.biopsych.2011.05.017 PubMed PMID: 21784414; PubMed Central PMCID: PMC3496792.

55. Chopra S, Segal A, Oldham S, Holmes A, Sabaroedin K, Orchard ER, et al. Network-Based Spreading of Gray Matter Changes Across Different Stages of Psychosis. JAMA Psychiatry. 2023 Dec 1;80(12):1246–57. doi:10.1001/jamapsychiatry.2023.3293 PubMed PMID: 37728918; PubMed Central PMCID: PMC10512169.

56. Chopra S, Francey SM, O’Donoghue B, Sabaroedin K, Arnatkeviciute A, Cropley V, et al. Functional Connectivity in Antipsychotic-Treated and Antipsychotic-Naive Patients With First-Episode Psychosis and Low Risk of Self-harm or Aggression: A Secondary Analysis of a Randomized Clinical Trial. JAMA Psychiatry. 2021 Sep 1;78(9):994–1004. doi:10.1001/jamapsychiatry.2021.1422 PubMed PMID: 34160595; PubMed Central PMCID: PMC8223142.

