## Supplement for "Longitudinal Brain Correlates of Cognitive Performance in Early Psychosis"

**Supplemental Materials**

**Methods**

***Participants***

The Human Connectome Project for Early Psychosis (HCP-EP) is a cross-sectional case-control study studying early psychosis across 4 sites in the United States. A total of 125 psychosis participants (91 non-affective and 34 affective) and 58 matched healthy controls completed study measures and were released in the September 2020 HCP-EP Release 1.1 on the National Data Archive (Supplemental Table 1, Figure 4). The study protocols were approved by the institutional review boards of Indiana University, Indianapolis, Indiana; Beth Israel Deaconess Medical Center, Boston, Massachusetts; and the Partners Healthcare IRB (now MGB sIRG) for Brigham and Women’s Hospital, Boston, Massachusetts, McLean Hospital, Belmont, Massachusetts, and Massachusetts General Hospital, Boston, Massachusetts. An additional 142 participants (150 early psychosis, 26 healthy controls) were enrolled at the Boston sites (R01 MH117012). All participants provided written informed consent. Longitudinal assessments were collected at the Boston sites only.

***Inclusion and Exclusion Criteria***

Medically stable male and female subjects with a confirmed psychiatric diagnosis and healthy control subjects were enrolled in the study. All individuals were 16-35 years of age at study entry, willing to share de-identified data with the Connectome database, and met inclusion criteria for one of the three subject cohorts. To be included in the Non-Affective Psychosis cohort, individuals had a DSM-V diagnosis of schizophrenia, schizophreniform, schizoaffective, psychosis not otherwise specified, delusional disorder, or brief psychotic disorder with onset within the past 5 years prior to study entry. Individuals in the Affective Psychosis cohort had a DSM-V diagnosis of major depression with psychosis (single and recurrent episodes) or bipolar disorder with psychosis (including most recent episode depressed and manic types) with onset within 5 years prior to study entry. Healthy controls were not permitted to meet criteria for bipolar and related disorders, major depressive disorder (recurrent), schizophrenia and other psychotic disorders, a current anxiety disorder. Healthy controls with a history of anxiety disorder were permitted in the study if the total duration of illness was less than 12 months, was in remission for at least 12 months, and did not require use of medication. Healthy controls were not permitted to have any first-degree family member diagnosed with a schizophrenia spectrum disorder, be taking any psychiatric medications at time of study entry, or have any history of psychiatric hospitalization. Individuals were excluded from any cohort if they had a substance-induced psychosis or psychotic disorder due to a medical condition, known IQ < 70 based on medical history, known medical history of HIV, an active medical condition that affected brain or cognitive functioning (e.g. seizure disorder, epilepsy, head trauma, stroke, traumatic brain injury, significant loss of consciousness, or other neurological disorder) in the site principal investigator’s opinion, implanted pacemaker, medication pump, vagal stimulator, deep brain stimulator, TENS unit, ventriculoperitoneal shunt, or other contraindication to undergoing an MRI scan, current severe substance use disorder in past 90 days (excluding caffeine and nicotine), electroconvulsive therapy treatment in past 12 months, high risk for suicidal acts, including active suicidal ideation or any suicide attempt within 30 days prior to screening, aggressive behavior or risk of substantial danger.

***Measures***

The Structured Clinical Interview for DSM-5-RV was used in conjunction with medical records and/or clinical interviews was administered to all subjects, both patients and healthy controls, to rule out patient subjects who were not psychotic or who had a psychosis that was related to substance abuse or an organic disease. The SCID-5-RV was also used to confirm that healthy control subjects did not meet criteria for bipolar and related disorders, major depressive disorder (recurrent), or schizophrenia and other psychotic disorders. Lifetime medical history was assessed during screening, including current medications.

**Auditory Continuous Performance Task (ACPT)**

Participants performed the Auditory Continuous Performance Test (ACPT) (1, 2). The ACPT is comprised of 4 task conditions which differ based on degree of working memory and interference load. For each task condition, letters of the alphabet are presented monaurally at a rate of one letter per second for four blocks of 90 seconds. Subjects are required to respond to all target stimuli by lifting their index finger.

The simplest target vigilance condition (ACPT Vigilance) requires subjects to respond to each A only if immediately preceded by a Q (i.e. QA, instructions: *“Respond to A immediately following a Q.”*). Increased working memory load without interference is assessed in the ACPT Memory condition. In the target condition for the ACPT Memory task, subjects respond to each A when preceded by a Q and separated by three letters (e.g. Q b t r A, instructions: *“Respond to A when preceded by a Q four letters previously.”*), and there are never Qs or As between the Q (warning) and A (target) (i.e. no interference).

To make the task more difficult, combinations of the letters Q, A, or QA were periodically embedded between the warning Q and target A (i.e. Q Q c q A A b r ) (i.e. interference). In this example, capital Qs and As were cues and targets, respectively, whereas the lowercase q is a distracter. Trials with interspersed Qs and interleaved series were designed to produce distraction, divide attention, and prevent counting, as the subject was episodically required to maintain two separate tracks simultaneously (e.g. constant updating of identification of stimuli from memory). There are two levels of the ACPT Interference condition. In the Low Interference condition, which has a low working memory load and low interference load, subjects respond to each A when preceded by a Q and separated by two letters previously (q s Q a A c g a Q Q A A m s r ). In the High Interference condition, which has a high working memory load and a high interference load, subjects respond to each A when preceded by a Q and separated by four letters previously (q s Q b r a A c g Q z Q h A p A m ). In HCP-EP, the Low Interference condition was used, while in NAPLS2, the High Interference condition was used. We summed performance on all 3 conditions of the ACPT to calculate a total score that reflects overall performance on the ACPT. ACPT total scores range from 0 to 300, with a higher number indicating better performance.

***Imaging Analysis***

All participants underwent a 5.6 min eyes-open resting-state scan (420 whole-brain volumes) where subjects were instructed to remain still during scanning, and deformable foam cushioning was used to stabilize the head. Real time image reconstruction and processing were used for quality assurance at the time of scanning. If there were any detectable problem, the scan was repeated. Noise-attenuating headphones and ear stopples were used and provided excellent noise reduction while still permitting adequate auditory perception. Data were acquired from three Siemens MAGNETOM Prisma 3T scanners at Brigham & Women’s Hospital (BWH), McLean Hospital, and Indiana University (IU). BWH and IU used a 32-channel head coil. McLean used a 64-channel head and neck coil, with the neck channels turned off. All protocols were based on the 2016 CCF template protocol. Functional images were collected using the following parameters: 2mm isotropic resolution, multiband acceleration factor of 8, TR 800ms, TE 37ms, 52 degree flip angle, 72 2-mm slices, 208-mm FOV acquired twice: one with AP and once with PA phase encoding. For this analysis, only the PA phase-encoded direction was used. In addition, high-resolution T1-weighted images were acquired for each participant with the following parameters: 0.8mm isotropic resolution, 256mm FOV, TR 2400ms, TE 2.22ms, 8 degree flip angle.

***MRI Data Processing***

All analyses were preprocessed using the DPABI toolbox (Data Processing and Analysis for Brain Imaging (3); http:// rfmri.org/dpabi). As a quality control metric, data from any participant whose scans exceeded motion thresholds (3 mm translation or 3° rotation) were discarded. Individual time points with framewise displacement 0.2 mm were removed via scrubbing (4), and scans with 50% of volumes removed for framewise displacement were discarded. All data were preprocessed to remove motion (24-parameter), CSF signals, white matter signals, global signal, and overall linear trend. A bandpass filter was applied (0.01–0.08 Hz). Data were normalized using the DARTEL toolbox into Montreal Neurological Institute (MNI) space and smoothed with an 8-mm full-width half-maximum kernel. Analyses were conducted in a gray matter mask defined within the group. Data were resampled into 4mm isotropic resolution.

*Seed-Based Connectivity Analysis:*

To calculate prefrontal-somatomotor connectivity, the time course of the BOLD signals from rsfMRI scans was extracted from the previously identified prefrontal region and a 6mm sphere (seed) placed at the previously identified location of maximal connectivity-cognition association in the somatomotor cortex (MNI X= +4, Y = -40, Z = +68) (5). We then modeled the association between prefrontal-somatomotor connectivity with ACPT total score.

**Supplemental Tables**

**Supplemental Table 1**

| **Variable** | **Estimate** | **SE** | **df** | **t** | **p** |
| --- | --- | --- | --- | --- | --- |
| Intercept | 272.04 | 3.78 | 331.04 | 72.06 | **<.0001** |
| Time from Baseline | 0.021 | 0.0051 | 277.40 | 4.09 | **<.0001** |
| Psychosis | -24.96 | 4.29 | 307.39 | -5.81 | **<.0001** |

**Supplemental Table 2**

| **Variable** | **Estimate** | **SE** | **df** | **t** | **p** |
| --- | --- | --- | --- | --- | --- |
| Intercept | 272.98 | 3.94 | 354.43 | 69.26 | **<.0001** |
| Time from Baseline | 0.015 | 0.0089 | 401.02 | 1.66 | 0.098 |
| Psychosis | -26.20 | 4.55 | 351.70 | -5.76 | **<.0001** |
| Time from Baseline*Psychosis | 0.0088 | 0.011 | 346.50 | 0.81 | 0.42 |

**Supplemental Table 3**

| **Variable** | **Estimate** | **SE** | **df** | **t** | **p** |
| --- | --- | --- | --- | --- | --- |
| Intercept | 237.70 | 3.05 | 241.40 | 77.97 | **<.0001** |
| Time from Baseline | 0.020 | 0.0068 | 173.80 | 2.94 | **0.0037** |
| Affective Psychosis | 22.55 | 4.70 | 215.70 | 4.80 | **<.0001** |

**Supplemental Table 4**

| **Variable** | **Estimate** | **SE** | **df** | **t** | **p** |
| --- | --- | --- | --- | --- | --- |
| Intercept | 236.68 | 3.09 | 254.43 | 76.65 | **<.0001** |
| Time from Baseline | 0.035 | 0.011 | 199.13 | 3.21 | **0.0015** |
| Affective Psychosis | 25.17 | 4.91 | 253.71 | 5.13 | **<.0001** |
| Time from Baseline*Affective Psychosis | -0.024 | 0.014 | 182.93 | -1.75 | 0.082 |

**Supplemental Table 5**

| **Variable** | **Estimate** | **SE** | **df** | **t** | **p** |
| --- | --- | --- | --- | --- | --- |
| Intercept | 235.83 | 5.80 | 163.78 | 40.64 | **<.0001** |
| DMPFC-Somatomotor Connectivity | 12.56 | 10.00 | 61.31 | 1.26 | 0.21 |
| Affective Psychosis | 28.82 | 8.33 | 137.76 | 3.46 | **<.0001** |
| Time from Baseline | 0.021 | 0.0070 | 35.47 | 3.02 | **0.0047** |
| Antipsychotic Dose (CPZeq) | -0.032 | 0.013 | 87.59 | -2.42 | **0.017** |

CPZeq: chlorpromazine equivalent dose (mg).

**Supplemental Table 6**

| **Variable** | **Estimate** | **SE** | **df** | **t** | **p** |
| --- | --- | --- | --- | --- | --- |
| Intercept | 242.04 | 6.05 | 164.69 | 40.01 | **<.0001** |
| DMPFC-Somatomotor Connectivity | 47.67 | 15.89 | 100.98 | 3.00 | **0.0034** |
| Affective Psychosis | 19.11 | 8.78 | 159.79 | 2.18 | **0.031** |
| Time from Baseline | 0.020 | 0.0072 | 33.91 | 2.84 | **0.0077** |
| Antipsychotic Dose (CPZeq) | -0.029 | 0.013 | 96.88 | -2.17 | **0.033** |
| DMPFC-Somatomotor Connectivity*Affective Psychosis | -53.36 | 19.48 | 67.80 | -2.74 | **0.0079** |

CPZeq: chlorpromazine equivalent dose (mg).

**Supplemental Table 7**

| **Variable** | **Estimate** | **SE** | **df** | **t** | **p** |
| --- | --- | --- | --- | --- | --- |
| Non-Affective | 47.67 | 16.70 | 120.50 | 2.86 | 0.0051 |
| Affective Psychosis | -5.69 | 13.10 | 58.20 | -0.43 | 0.67 |

**Supplemental Table 8**

| **Variable** | **Estimate** | **SE** | **df** | **t** | **p** |
| --- | --- | --- | --- | --- | --- |
| Intercept | -0.42e | 0.14 | 129.50 | -3.05 | 0.0028 |
| Fluid Cognition | -0.0011 | 0.0014 | 110.10 | -0.81 | 0.42 |
| ACPT Total | 0.0015 | 0.00060 | 127.90 | 2.41 | **0.017** |
| Affective Psychosis | -0.0028 | 0.049 | 95.43 | -0.56 | 0.57 |
| Time from Baseline | -0.00016 | 0.000089 | 104.50 | -1.75 | 0.083 |
| Antipsychotic Dose (CPZeq) | -0.000058 | 0.00011 | 133.60 | -0.55 | 0.58 |

CPZeq: chlorpromazine equivalent dose (mg).

**Supplemental Table 9**

| **Variable** | **Estimate** | **SE** | **df** | **t** | **p** |
| --- | --- | --- | --- | --- | --- |
| Intercept | -0.48 | 0.14 | 130.40 | -3.38 | 0.00095 |
| Fluid Cognition | -0.0013 | 0.0014 | 108.80 | -0.93 | 0.36 |
| ACPT Total | 0.0018 | 0.00063 | 128.20 | 2.80 | **0.0059** |
| Affective Psychosis | 0.52 | 0.36 | 122.90 | 1.44 | 0.15 |
| Time from Baseline | -0.00013 | 0.000090 | 110.40 | -1.46 | 0.15 |
| Antipsychotic Dose (CPZeq) | -0.000064 | 0.00011 | 133.30 | -0.61 | 0.55 |
| ACPT Total*Affective Psychosis | -0.0021 | 0.0014 | 122.40 | -1.53 | 0.13 |

CPZeq: chlorpromazine equivalent dose (mg).

**References**

1.	Seidman LJ, Breiter HC, Goodman JM, Goldstein JM, Woodruff PW, O'Craven K, et al. A functional magnetic resonance imaging study of auditory vigilance with low and high information processing demands. Neuropsychology. 1998;12(4):505–18.

2.	Seidman LJ, Meyer EC, Giuliano AJ, Breiter HC, Goldstein JM, Kremen WS, et al. Auditory working memory impairments in individuals at familial high risk for schizophrenia. Neuropsychology. 2012;26(3):288–303.

3.	Yan CG, Wang XD, Zuo XN, Zang YF. DPABI: Data Processing & Analysis for (Resting-State) Brain Imaging. Neuroinformatics. 2016;14(3):339–51.

4.	Power JD, Barnes KA, Snyder AZ, Schlaggar BL, Petersen SE. Spurious but systematic correlations in functional connectivity MRI networks arise from subject motion. Neuroimage. 2012;59(3):2142–54.

5.	Ward HB, Beermann A, Xie J, Yildiz G, Felix KM, Addington J, et al. Robust Brain Correlates of Cognitive Performance in Psychosis and Its Prodrome. Biol Psychiatry. 2025;97(2):139–47.
